# A causal association between schizophrenia and bipolar disorder on rheumatoid arthritis: A two-sample Mendelian randomization study

**DOI:** 10.1101/2021.08.12.21261493

**Authors:** Gonul Hazal Koc, Fatih Ozel, Kaan Okay, Dogukan Koc, Pascal H.P. de Jong

## Abstract

**Background:** Schizophrenia (SCZ) and bipolar disorder (BD) are both associated with several autoimmune disorders including rheumatoid arthritis(RA). However, a causal association of SCZ and BD on RA is controversial and elusive. In the present study, we aimed to investigate the causal association of SCZ and BD with RA by using the Mendelian randomization (MR) approach.

**Methods:** A two-sample MR (2SMR) study including the inverse-variance weighted(IVW), weighted median, simple mode, weighted mode and MR-Egger methods were performed. We used summary-level genome-wide association study(GWAS) data in which BD and SCZ are the exposure and RA the outcome. We used data from the Psychiatric Genomics Consortium(PGC) for BD(n= 41,917) and SCZ(n= 33,426) and RA GWAS dataset(n= 2,843) from the European ancestry for RA.

**Results:** We found 48 and 52 independent single nucleotide polymorphisms (SNPs, r2 <0.001)) that were significant for respectively BD and SCZ (p <5×10-8). Subsequently, these SNPs were utilized as instrumental variables(IVs) in 2SMR analysis to explore the causality of BD and SCZ on RA. The two out of five MR methods showed a statistically significant inverse causal association between BD and RA: weighted median method(odds ratio (OR), 0.869, [95% CI, 0.764-0.989]; *P*= 0.034) and inverse-variance weighted(IVW) method (OR, 0.810, [95% CI, 0.689-0.953]; *P*= 0.011). However, we did not find any significant association of SCZ with RA (OR, 1.008, [95% CI, 0.931-1.092]; *P*= 0.829, using the IVW method).

**Conclusions:** These results provide support for an inverse causal association between BD and RA. Further investigation is needed to explain the underlying protective mechanisms in the development of RA.

**Key messages:**

- Mendelian randomization can offer strong insight into the cause-effect relationships in rheumatology.
- Bipolar disorder had a protective effect on rheumatoid arthritis.
- There is no inverse causal association between schizophrenia and rheumatoid arthritis contrary to the findings from observational studies.

## Introduction

Schizophrenia (SCZ) and bipolar disorder (BD) are severe psychiatric disorders that are characterized by significant emotional, cognitive and behavioral symptoms, and they cause substantial deterioration in functioning [1, 2]. Both disorders pose an increased risk of morbidity and mortality and are associated with decreased life expectancy [3-5]. Genetic factors have been investigated for SCZ [6] and the interplay between genetic and environment has also been a focus [7, 8]. Still, the etiopathogenesis of SCZ seems heterogeneous, and the existing evidence remains inconclusive. Many risk factors have also been identified for BD in a similar manner [9]. However, our understanding of the biological underpinnings is limited. Notwithstanding, disturbances in inflammatory signaling mechanisms seem to play a critical role in the etiopathogenesis of SCZ and BD [10, 11]. For example, several studies have shown that autoimmune disorders are more prevalent in SCZ and BD compared to the general population, but potential confounders were not always addressed correctly [12, 13].

Rheumatoid arthritis (RA) on the other hand is a common autoimmune disorders that is caused by an dysregulation of the immune system [14]. The disease may cause severe function impairment [15]. RA is also accompanied with several comorbidities, that can have a severe impact on patients’ lives. One of these comorbidities might be psychiatric disorders on which we will focus.

Population-based studies mainly demonstrated the lower prevalence of RA in patients with SCZ [16-18]. The possible explanations for the lower prevalence of RA and SCZ have not been examined comprehensively, and the lack of evidence raises the demand for exploring this relationship with various potent methods. On the other hand, the high comorbidity of RA has been speculated for BD in the literature. Two different case-control studies demonstrated a higher prevalence of RA among patients with BD [19, 20]. The increased risk of developing BD in persons with RA has also been established in population-based settings [21-23]. Even though, a high prevalence of BD among patients with RA has been shown in another case-control study [24], the association between BD and RA was not significant in the multivariate analysis, which posits the importance of potential confounding.

Although aforementioned findings suggest that the prevalence of RA is lower in SCZ and higher in BD, the evidence is still limited and the causal relationships are yet to be established. Nonetheless, inflammatory disturbances seem to be the main reason for the associations between these psychiatric disorders and RA; potential confounders need to be investigated thoroughly, to avoid biased associations. Regrettably, observational studies are limited to capture many confounders, and experimental designs carry other difficulties to conduct to scrutinize the co-existence of RA and SCZ or BD. Mendelian randomization (MR) may be a favorable solution to circumvent these shortcomings and explores the causation of a relationship between exposure and an outcome using genetic variants. MR is a robust epidemiological method that uses the genetic variants related to the exposure of interest to make causal inferences about the non-genetic measures of the outcome [25]. MR exploits single-nucleotide polymorphisms (SNPs) as instrumental variables (IVs) for the exposure [26]; thus, it can evade the issues caused by confounding and provide more accurate causal interpretations. MR can be a cost-effective solution for clinical studies that might otherwise be extremely costly, technically or ethically difficult. To our best knowledge, no MR study has been performed previously to explore the causality between RA and SCZ or BD. Therefore, our aim is to investigate the potential causal relationship between having SCZ or BD and the risk of developing RA.

## Materials and methods

We performed a two-sample mendelian randomization (2SMR) approach to assess causality between BD, SCZ, and RA using publicly available GWAS summary datasets.

### Data sources

We downloaded the latest GWAS summary data with BD I and II, which includes 7,608,183 single nucleotide polymorphisms (SNPs) of 41,917 cases and 371,549 controls [27] and the SCZ GWAS data including 8,379,106 SNPs of 33,426 cases and 32,541 controls [28] from the Psychiatric Genomics Consortium database (https://www.med.unc.edu/pgc/). The RA GWAS summary data included 8,514,610 SNPs of 2,843 cases and 5,540 controls [29] and was downloaded from the Japanese ENcyclopedia of GEnetic associations by Riken database (http://jenger.riken.jp/en/result). All GWAS summary data are based on European ancestry.

### Instrument identification

We used the R package TwoSampleMR v0.5.6 to harmonize SNP information and conducted a 2SMR analysis between BD, SCZ, and RA [30]. All GWAS data had beta coefficient (β), standard error of β (SE), major and minor alleles for each SNP together with the allele frequencies, *p*-value for relevant association, and total sample size information. We followed four steps in the analysis: *i)* genome-wide significant SNPs (*p* ≤ 5×10^−8^) related to exposure were selected as (IVs) (IVs); *ii)* linkage disequilibrium (LD) pruning (r^2^ <0.001) was performed to obtain independent SNPs; *iii)* genome-wide significant and independent SNPs were extracted from outcome GWAS data and those SNPs were utilized in harmonization of exposure and outcome GWAS data to ensure their effect on those data to correspond to the same allele; *iv)* harmonized data were used in a 2SMR analysis. Eventually, we identified 48 and 52 independent SNPs at genome-wide significance from respectively BD and SCZ GWAS data. These SNPs were used as IVs for harmonization, MR, heterogeneity, and sensitivity analyses in the R computation environment v4.1.0 (http://www.R-project.org).

### Two-sample Mendelian randomization

The β and SE were used in the estimation of the causal relationship. We obtained MR results for the following five methods: Inverse-variance weighted (IVW) [31], MR-Egger [32], weighted median [33], weighted mode and simple mode [34]. Either BD or SCZ were the exposure and RA was for both exposures the outcome. The respectively 48 and 52 independent SNPs were used as IVs in the 2SMR analysis. Subsequently, the aforementioned five MR methods were utilized to generate effect size estimates of these SNPs.

### Sensitivity analyses

Sensitivity analyses were performed to examine the existence of horizontal pleiotropy and heterogeneity. In the case of horizontal pleiotropy, a single locus can affect an outcome through one or more biological pathways independent of that of the assumed exposure. To test the horizontal pleiotropic effect amongst BD, SCZ, and RA, we performed MR-Egger regression analysis. Furthermore, we employed the leave-one-out analysis, where one variant that is strongly associated with exposure and dominates the estimate of the causal effect is removed from the analysis to re-estimate the causal effect.

## Results

After genome-wide significance filtering, LD pruning, and harmonization of potential SNPs, 48 BD and 52 SCZ independent genome-wide significant SNPs were considered as IVs. These IVs were used in a 2SMR analysis to estimate the causality of BD and SCZ on RA. The detailed information on these IVs can be found in **Supplementary Table S1** and **S2**. The relationship is represented as odds ratios (ORs) with respective p-values in **Table 1**.

**Table 1.**
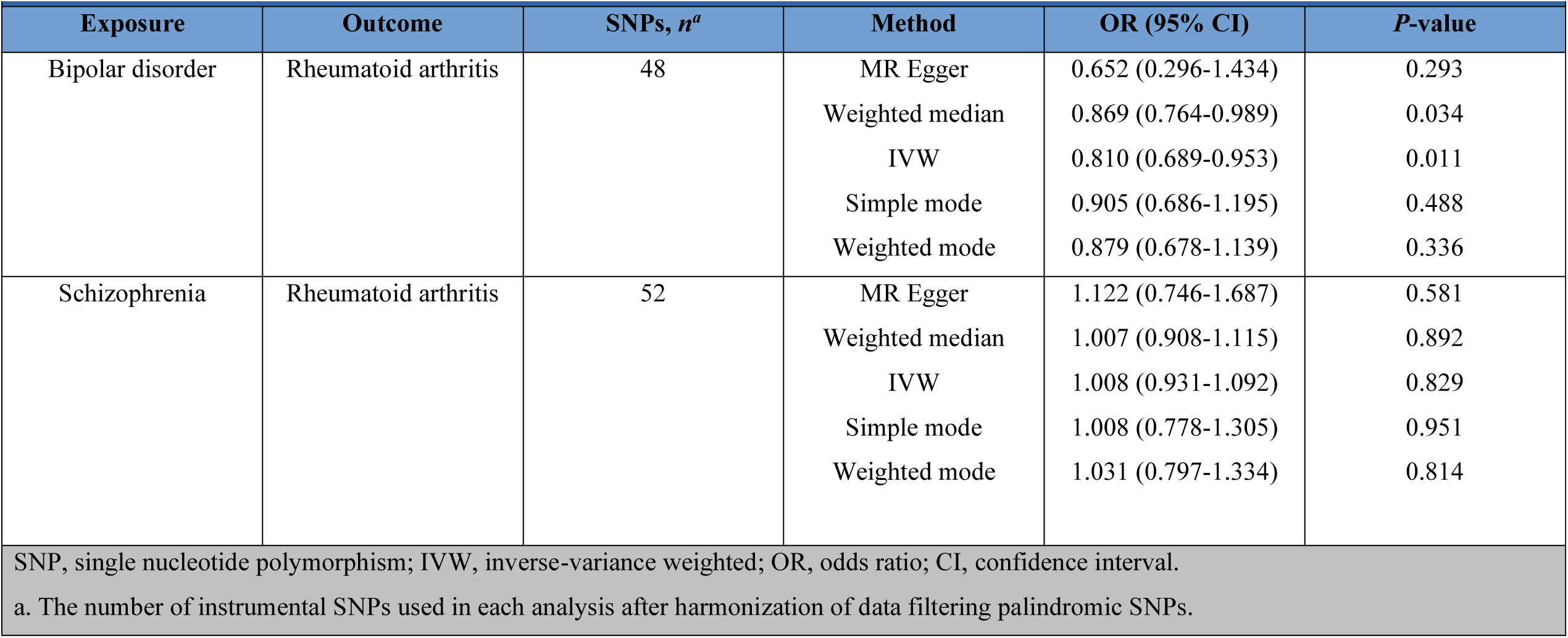
Two-sample Mendelian Randomization analysis of the effects of bipolar disorder, schizophrenia, and rheumatoid arthritis using summary-level data.

| Exposure | Outcome | SNPs, *n*<sup>a</sup> | Method | OR (95% CI) | *P*-value |
| --- | --- | --- | --- | --- | --- |
| Bipolar disorder | Rheumatoid arthritis | 48 | MR Egger | 0.652 (0.296-1.434) | 0.293 |
|  |  |  | Weighted median | 0.869 (0.764-0.989) | 0.034 |
|  |  |  | IVW | 0.810 (0.689-0.953) | 0.011 |
|  |  |  | Simple mode | 0.905 (0.686-1.195) | 0.488 |
|  |  |  | Weighted mode | 0.879 (0.678-1.139) | 0.336 |
| Schizophrenia | Rheumatoid arthritis | 52 | MR Egger | 1.122 (0.746-1.687) | 0.581 |
|  |  |  | Weighted median | 1.007 (0.908-1.115) | 0.892 |
|  |  |  | IVW | 1.008 (0.931-1.092) | 0.829 |
|  |  |  | Simple mode | 1.008 (0.778-1.305) | 0.951 |
|  |  |  | Weighted mode | 1.031 (0.797-1.334) | 0.814 |

SNP, single nucleotide polymorphism; IVW, inverse-variance weighted; OR, odds ratio; CI, confidence interval.
a. The number of instrumental SNPs used in each analysis after harmonization of data filtering palindromic SNPs.

We used five MR methods to estimate the causality of BD and SCZ on RA. We found an inverse causal association between BD and RA. MR methods weighted median (OR= 0.869, [95% CI 0.764-0.989]; *P*= 0.034) and IVW (OR= 0.810, [95% CI 0.689-0.953]; *P*= 0.011) were significant, while the other methods were not (MR Egger (OR= 0.652, [95% CI 0.296-1.434]; *P*= 0.293), simple mode (OR= 0.905, [95% CI 0.686-1.195]; *P*= 0.488), and weighted mode (OR= 0.879, [95% CI 0.678-1.139]; *P*= 0.336). These estimates are available in **Figure 1A** and **Table 1** in detail. We also performed a leave-one-out analysis and did not find a notable effect of any single SNP that could dominate the results (**Supplementary Figure S1**). The funnel plot did not signify that there were heterozygous SNPs (**Figure 1B**). We did not find evidence for pleiotropy from the MR-Egger regression (intercept: 0.015, *P*= 0.583). The heterogeneity results can be found in **Supplementary Table S3**. On the other hand, no statistical significance in the causal relationship between SCZ and RA was found (**Table 1**). More detailed results of these estimations are presented as respectively scatter and funnel plots in **Figure 1C-D**.

**Figure 1.**
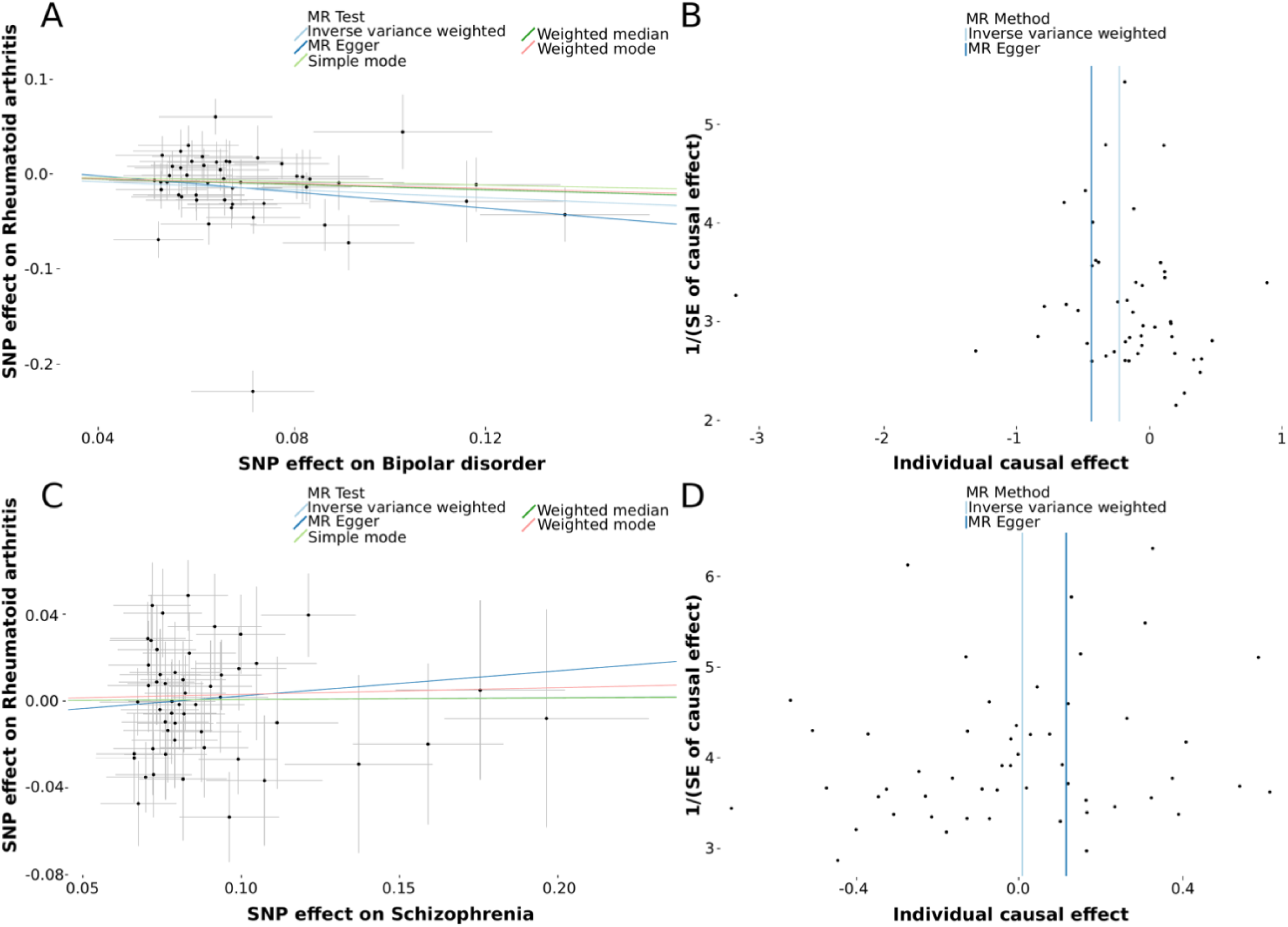
**A**. Scatter plot of single nucleotide polymorphism (SNP) potential effects on bipolar disorder versus rheumatoid arthritis. The 95% CI for the effect size on rheumatoid arthritis is shown as vertical lines, while the 95% CI for the effect size on bipolar disorder is shown as horizontal lines. The slope of fitted lines represents the estimated Mendelian randomization effect per method. **B**. Funnel plot for bipolar disorder shows the estimation using the inverse of the standard error of the causal estimate with each individual SNP as a tool. The vertical line represents the estimated causal effect obtained using IVW and MR-Egger methods. **C**. Scatter plot of SNP potential effects on schizophrenia versus rheumatoid arthritis **D**. Funnel plot for schizophrenia shows the estimation using the inverse of the standard error of the causal estimate with each individual SNP as a tool.

## Discussion

We performed 2SMR analyses to explore the causality between SCZ, BD and RA based on summary statistics of the largest available GWAS to date. Our analyses suggested that BD had a protective effect on RA, whereas SCZ had no significant effect on RA. Considering the power of the MR approach on the exclusion of confounding factors, the effects of environmental factors can be decreased because genetic variants are assigned randomly at conception, similar to randomization in a clinical trial. From this standpoint, our study can be considered as the first potential evidence for uncovering causal relationships between psychiatric disorders (SCZ, BD) and RA.

A large number of existing studies in the broader literature have shown an inverse association between SCZ and RA [17, 18, 35-38]. It has been hypothesized that SCZ is a protective factor for RA, some focusing on environmental exposures and immunity, others on misclassification biased by underdiagnosis of RA in patients with SCZ [18, 39, 40]. A recent population-based case-control study from Sweden demonstrated that patients with SCZ have a decreased risk of developing RA (hazard ratio [HR] = 0.69, 95% CI = 0.59–0.80) [18].Similar results were found in the Danish Psychiatric Case Register study involving 20,495 patients with SCZ and 204,912 randomly selected age- and sex-matched controls (HR= 0.44, 95% CI = 0.24–0.81) [17]. Our finding contradict aforementioned studies. A possible explanation might be that causal evidence from observational studies are effected by (un)known confounding factors. For example, the mean age of onset for RA is usually later in life than SCZ and comorbid medical conditions are tend to be underreported in patients with SCZ [18, 19], which might be the case of the found decreased risk in these case-control studies. Therefore, it is important that screening for comorbidities in SCZ patients should be done more thoroughly, which is also supported by our results.

Various clinical studies have been conducted in the past to study the co-occurrence of BD and RA [19, 21, 41]. Nevertheless, the association between BD and RA remained unclear and debatable. For example, a Swedish register study in a sample of 3,798 patients with BD and 6,485 controls showed a greater risk for RA in BD compared to controls [19]. Similar results were found in a Danish and Taiwan registry [21, 41]. On the other hand, a large Swedish Population Registry showed a similar incidence of RA in patients with BD [18]. In this registry 34,744 patients with BD were included, who had a relative risk of developing RA of 1.01 (0.96 – 1.06, 95% CI). Moreover, a nationally representative longitudinal study from the Netherlands showed that having any mood or any anxiety disorder did not predict new-onset arthritis [42]. However, several inevitable confounding factors in observational studies are not adequately addressed in the literature. In the present study, we aimed to clarify the causal association between BD and RA. Although our analysis showed that patients with BD had a lower chance of developing RA, the reasons behind this protective effect are yet unclear. Fortunately, several studies already investigated the relationship between autoimmune disorders and severe psychiatric conditions, including BD and SCZ [43-46]. Immunity and gene-environment interactions represent a major risk factor for these disorders [47-50]. The studies showed that the major histocompatibility complex (MHC) was one of the best validated genetic susceptibility loci for major psychiatric and autoimmune disorders [44, 51, 52]. A growing body of evidence also suggested that MHC involvement is more prominent in SCZ compared to BD [51]. Interestingly, some MHC loci have been revealed to contribute predisposition to certain autoimmune conditions while others are protective [44]. Therefore, it is important to highlight the fact that the prevalence of comorbid autoimmune conditions in BD and SCZ differ significantly, which is also seen in our results [19].

The present study has several strengths. We used the latest accessible and analyzable BD GWAS data providing confidence in MR assumptions to estimate the causality. All participants in the GWAS data were of European ancestry, which facilitated the elimination of confounding factors stemming from different races. Furthermore, we observed no horizontal pleiotropy, that is, the outcome directly affected by IVs of exposure without confounding. This provides robust causality of BD on RA and eases the interpretation of the genetic basis of the relationship. A potential limitation of our study is that in the direction of BD to RA, effect size estimates had borderline significance, which could be due to relatively limited sample size and/or low causal relationship. Also, we postulated a linear relationship between exposure and outcome, while using summary data that excluded other interactions (e.g., gene-gene and gene-environment). This might have led to deviations from this linearity. Finally, the clumping algorithm that identified the correlated genetic variants depended solely on European samples, because of the lack of corresponding data from other populations. This makes it hard to generalize the data to other populations (e.g. Asian).

In conclusion, we found no causal association between SCZ and RA, which contradicts current knowledge, and therefore we advise to screen SCZ more thoroughly for joint complaints. On the other hand the risk of RA is reduced in BD, however future studies are necessary to explore the biological mechanisms underlying this genetically predicted relationship.

## Data Availability

Only publicly available data were used in this study. Data sources and handling of these data are described in the Materials and methods. The codes can be found in R package TwoSampleMR.

## Ethics approval and consent to participate

The present study used only publicly available summary-level statistics. No individual-level data was analyzed. Ethical approval is therefore not required.

## Acknowledgements

The authors thank all individuals who shared GWAS summary statistics.

