## Supplementary material for "A causal association between schizophrenia and bipolar disorder on rheumatoid arthritis: A two-sample Mendelian randomization study" (from another version of this preprint: version 1, 2021-08-13)

| Supplementary Table S1. SNPs selected as instrumental variables and their associations with bipolar disorder. | | | | | | | | |
|---|---|---|---|---|---|---|---|---|
| SNP | Chromosome | Position (hg19) | Gene | Annotation | Effect Allele | Other Allele | β | se (β) |
| rs10043984 | 5 | 137712121 | KDM3B | intron | C | T | -0.059 | 0.010 |
| rs10255167 | 7 | 140676153 | CCT4P1 | intergenic | G | A | -0.066 | 0.011 |
| rs10737496 | 1 | 163745389 | RP4-640E24.1 | intergenic | C | T | 0.054 | 0.009 |
| rs10791849 | 11 | 65890597 | PACS1 | intron | T | A | 0.069 | 0.012 |
| rs10866641 | 5 | 169289206 | DOCK2 | intron | T | C | 0.062 | 0.009 |
| rs10994415 | 10 | 62322034 | ANK3 | intron | T | C | -0.118 | 0.017 |
| rs11062170 | 12 | 2348844 | CACNA1C | intron | G | C | -0.077 | 0.009 |
| rs11764361 | 7 | 105043229 | SRPK2 | upstream | A | G | 0.061 | 0.010 |
| rs12575685 | 11 | 70517927 | SHANK2 | intron | G | A | -0.065 | 0.010 |
| rs12668848 | 7 | 2020995 | MAD1L1 | intron | G | A | 0.057 | 0.009 |
| rs12932628 | 16 | 89632725 | - | - | G | T | -0.057 | 0.009 |

| | | | | | | | | |
|---|---|---|---|---|---|---|---|---|
| rs13044225 | 20 | 60865815 | OSBPL2 | intron | A | G | -0.054 | 0.009 |
| rs13195402 | 6 | 26463575 | - | - | G | T | 0.136 | 0.017 |
| rs13417268 | 2 | 169481837 | CERS6 | intron | C | G | 0.061 | 0.011 |
| rs1487445 | 6 | 98565211 | RP11-436D23.1 | intron | C | T | -0.074 | 0.009 |
| rs17183814 | 2 | 166152389 | SCN2A | splice acceptor | G | A | 0.102 | 0.018 |
| rs174592 | 11 | 61618608 | FADS2 | intron | A | G | -0.072 | 0.009 |
| rs1998820 | 10 | 18751103 | CACNB2 | intron | T | A | 0.083 | 0.015 |
| rs2011302 | 2 | 193738336 | PCGEM1 | intergenic | T | A | -0.052 | 0.009 |
| rs2126180 | 1 | 61105668 | LINC01748 | intron | G | A | -0.056 | 0.009 |
| rs2273738 | 10 | 111648659 | XPNPEP1 | intron | C | T | -0.091 | 0.013 |
| rs228768 | 17 | 42191893 | HDAC5 | intron | G | T | 0.064 | 0.010 |
| rs2336147 | 3 | 52626443 | PBRM1 | intron | T | C | 0.067 | 0.009 |
| rs237460 | 20 | 48033127 | KCNB1 | intron | C | T | -0.055 | 0.009 |
| rs2693698 | 14 | 99719219 | BCL11B | intron | A | G | -0.053 | 0.009 |

| | | | | | | | | |
|---|---|---|---|---|---|---|---|---|
| rs28455634 | 16 | 9230816 | C16orf72 | intergenic | G | A | 0.062 | 0.009 |
| rs28565152 | 5 | 7542911 | ADCY2 | intron | G | A | -0.067 | 0.011 |
| rs2953928 | 8 | 34152492 | RP1-84O15.2 | intron | G | A | -0.116 | 0.02 |
| rs35306827 | 13 | 113869045 | CUL4A | intron | G | A | 0.065 | 0.011 |
| rs35958438 | 15 | 38973793 | RP11-275I4.2 | intron | G | A | 0.064 | 0.011 |
| rs41315395 | 6 | 32201469 | TBC1D22B | intergenic | C | A | -0.071 | 0.012 |
| rs4447398 | 15 | 42904904 | STARD9 | intron | A | C | 0.082 | 0.013 |
| rs4619651 | 2 | 97416153 | LMAN2L | intergenic | G | A | 0.066 | 0.010 |
| rs4790841 | 17 | 1835482 | RTN4RL1 | downstream | C | T | -0.072 | 0.013 |
| rs5758064 | 22 | 41153879 | SCUBE1 | intergenic | T | C | 0.052 | 0.009 |
| rs61554907 | 17 | 38220432 | THRA | intron | G | T | -0.086 | 0.015 |
| rs62489493 | 8 | 9763581 | LINC00599 | upstream | C | G | -0.089 | 0.013 |
| rs67712855 | 20 | 43682551 | STK4 | intron | T | G | 0.067 | 0.010 |
| rs6887473 | 5 | 80961069 | SSBP2 | intron | G | A | 0.060 | 0.010 |

| | | | | | | | | |
|---|---|---|---|---|---|---|---|---|
| rs6946056 | 7 | 131870597 | PLXNA4 | intron | A | C | -0.053 | 0.009 |
| rs6954854 | 7 | 21492589 | SP4 | intron | G | A | 0.058 | 0.009 |
| rs696366 | 3 | 107757060 | RP11-861A13.3 | noncoding exon | C | A | 0.051 | 0.009 |
| rs6992333 | 8 | 144993377 | PLEC | synonymous | A | G | -0.060 | 0.01 |
| rs7108878 | 11 | 79107807 | TENM4 | intron | T | G | -0.081 | 0.014 |
| rs7201930 | 16 | 9958655 | GRIN2A | intron | T | C | -0.058 | 0.010 |
| rs748455 | 15 | 85149575 | ZSCAN2 | intron | T | C | 0.067 | 0.010 |
| rs7707252 | 5 | 78840610 | CTD-2232E5.2 | intergenic | A | G | -0.057 | 0.010 |
| rs9834970 | 3 | 36856030 | LINC02033 | downstream | T | C | -0.083 | 0.009 |

SNP, single nucleotide polymorphism; OR, odds ratio; se, standard error; β, beta coefficient.

| Supplementary Table S2. SNPs selected as instrumental variables and their associations with schizophrenia. | | | | | | | | |
|---|---|---|---|---|---|---|---|---|
| **SNP** | **Chromosome** | **Position (hg19)** | **Gene** | **Annotation** | **Effect Allele** | **Other Allele** | **β** | **se (β)** |
| rs1024582 | 12 | 2402246 | CACNA1C | intron | A | G | 0.093 | 0.012 |
| rs11027827 | 11 | 24375179 | RP11-2F20.1 | intergenic | T | C | -0.073 | 0.012 |
| rs11210892 | 1 | 44100084 | PTPRF | intergenic | A | G | -0.083 | 0.012 |
| rs11602497 | 11 | 130717432 | RP11-890B15.2 | splice donor | A | G | -0.079 | 0.014 |
| rs11693094 | 2 | 185601420 | ZNF804A | intron | T | C | -0.080 | 0.012 |
| rs11740474 | 5 | 153680747 | GALNT10 | intron | A | T | -0.074 | 0.012 |
| rs1198588 | 1 | 98552832 | NFU1P2 | intergenic | A | T | -0.122 | 0.014 |
| rs12190758 | 6 | 93148341 | RP11-374I15.1 | intergenic | A | G | -0.076 | 0.013 |
| rs12421382 | 11 | 109378071 | RP11-708B6.2 | intron | T | C | -0.074 | 0.012 |
| rs12532143 | 7 | 111025948 | IMMP2L | intron | T | C | 0.079 | 0.012 |
| rs12704290 | 7 | 86427626 | GRM3 | intron | A | G | -0.107 | 0.018 |
| rs12826178 | 12 | 57622371 | SHMT2 | upstream | T | G | -0.175 | 0.026 |

| | | | | | | | | |
|---|---|---|---|---|---|---|---|---|
| rs13107325 | 4 | 103188709 | SLC39A8 | missense | T | C | 0.136 | 0.023 |
| rs13176930 | 5 | 152664721 | LINC01470 | intergenic | A | T | -0.071 | 0.012 |
| rs13262595 | 8 | 143316970 | TSNARE1 | intron | A | G | 0.070 | 0.012 |
| rs17620999 | 3 | 2561556 | CNTN4 | intron | A | G | -0.094 | 0.015 |
| rs1789589 | 18 | 53804156 | CTD-2008L17.2 | intron | T | C | -0.079 | 0.013 |
| rs2068012 | 14 | 30190316 | PRKD1 | intron | T | C | -0.080 | 0.014 |
| rs2103655 | 20 | 37425958 | RP11-101E14.3 | intergenic | A | G | 0.089 | 0.013 |
| rs2535627 | 3 | 52845105 | ITIH4 | downstream | T | C | 0.065 | 0.012 |
| rs281768 | 2 | 200825240 | MAIP1 | intron | A | T | 0.098 | 0.015 |
| rs2851447 | 12 | 123665113 | MPHOSPH9 | intron | C | G | -0.082 | 0.013 |
| rs34053174 | 14 | 99722290 | BCL11B | intron | C | G | -0.076 | 0.014 |
| rs34312605 | 3 | 180590297 | FXR1 | intron | T | C | -0.084 | 0.014 |
| rs34542576 | 14 | 71535493 | PCNX1 | intron | A | C | 0.069 | 0.012 |
| rs35017470 | 5 | 87727104 | LINC02060 | intron | T | C | 0.196 | 0.032 |
| rs3741434 | 12 | 53605344 | RARG | 3 prime UTR | T | C | -0.105 | 0.019 |
| rs3743078 | 15 | 78894759 | CHRNA3 | intron | C | G | -0.099 | 0.014 |

| | | | | | | | | |
|---|---|---|---|---|---|---|---|---|
| rs3802924 | 11 | 133827733 | IGSF9B | upstream | A | C | 0.090 | 0.014 |
| rs3849046 | 5 | 137851192 | ETF1 | intron | T | C | 0.066 | 0.012 |
| rs4702 | 15 | 91426560 | FURIN | 3 prime UTR | A | G | -0.087 | 0.012 |
| rs4766428 | 12 | 110723245 | ATP2A2 | intron | T | C | 0.071 | 0.012 |
| rs4906364 | 14 | 104214342 | PPP1R13B | intron | C | G | -0.082 | 0.012 |
| rs55661361 | 11 | 124613957 | NRGN | intron | A | G | -0.082 | 0.012 |
| rs61882743 | 11 | 46548754 | AMBRA1 | intron | C | G | -0.096 | 0.015 |
| rs6704768 | 2 | 233592501 | GIGYF2 | intron | A | G | -0.068 | 0.012 |
| rs67119537 | 15 | 84950369 | RP11-182J1.14 | - | T | C | 0.081 | 0.013 |
| rs6868457 | 5 | 60550041 | LINC02057 | intron | T | C | 0.075 | 0.012 |
| rs7085104 | 10 | 104628873 | AS3MT | upstream | A | G | 0.098 | 0.012 |
| rs7185124 | 16 | 13750694 | U95743.1 | intergenic | T | C | 0.088 | 0.014 |
| rs72936314 | 18 | 53580677 | LINC01416 | intron | A | G | 0.158 | 0.023 |
| rs7577690 | 2 | 220045035 | FAM134A | intron | A | G | 0.070 | 0.012 |
| rs7599488 | 2 | 60718347 | BCL11A | intron | T | C | -0.072 | 0.012 |
| rs760648 | 22 | 42571028 | TCF20 | intron | A | G | 0.076 | 0.013 |
| rs7653924 | 4 | 170202862 | LINC02174 | intergenic | T | C | 0.065 | 0.012 |

| | | | | | | | | |
|---|---|---|---|---|---|---|---|---|
| rs7819570 | 8 | 89588626 | RP11-586K2.1 | intron | T | G | 0.085 | 0.015 |
| rs7893279 | 10 | 18745105 | CACNB2 | intron | T | G | 0.110 | 0.019 |
| rs8139773 | 22 | 39975691 | CACNA1I | intron | A | G | -0.075 | 0.012 |
| rs832190 | 3 | 63842629 | THOC7 | intron | T | C | -0.073 | 0.012 |
| rs9330316 | 2 | 110284236 | SEPT10 | intergenic | A | G | 0.071 | 0.013 |
| rs940174 | 3 | 136309710 | STAG1 | intron | T | C | 0.071 | 0.012 |
| rs9428966 | 1 | 243667900 | AKT3 | 3 prime UTR | T | G | 0.077 | 0.013 |

SNP, single nucleotide polymorphism; OR, odds ratio; se, standard error; β, beta coefficient.

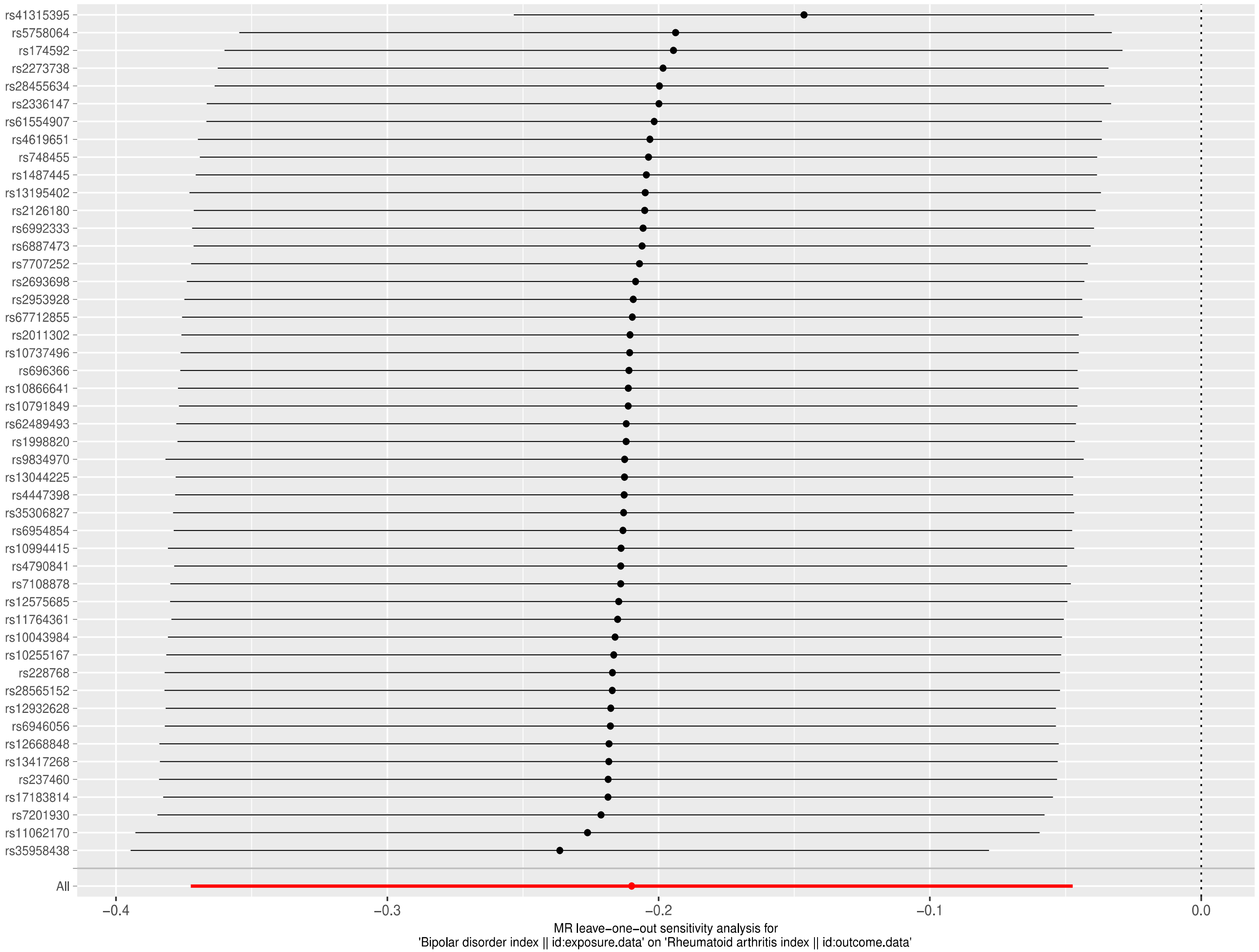


**Supplementary Figure S1.** Leave-one-out analysis for the estimates for bipolar disorder on rheumatoid arthritis. Each point represents the Mendelian randomization estimate excluding that particular variant. The leave-one-out plot suggests that the association determined is not disproportionately affected by a certain instrumenta variable.
